# Hepatitis E Virus Prevalence and Associated Risk Factors in Pregnant Women Attending Antenatal Consultations Niger Republic, 2025

**DOI:** 10.64898/2026.10.05.26364823

**Authors:** Fatimata Hassane, Adamou Lagare, Abdourahamane Yacouba, Hadiza Ousmane, Moussa Issa, Larwanou Harouna Magagi, Balki Aoula, Bintou Kiari Kaka, Oumarou Diallo, Médard Djedanem, Mahamadou Doucthi, Mamadou Saidou

**Affiliations:** Centre de Recherche Médicale et Sanitaire (CERMES), Niamey, Niger; Faculty of Health Sciences, University Abdou Moumouni, Niamey, Niger; Epicentre, Médecins sans frontières, Niamey, Niger; Faculty of Health Sciences, University André Salifou de Zinder, Zinder, Niger

**Keywords:** Hepatitis E virus, pregnant women, prevalence, risk factors

## Abstract

**Background:** Hepatitis E virus (HEV) is highly endemic in several African countries with high mortality rate among pregnant women. The prevalence of HEV in Niger among pregnant women is unknown. We hereby aim to report the prevalence and risk factors of HEV among pregnant women in Niamey, Niger.

**Methods:** Blood samples were collected from pregnant women attending antenatal consultation in three selected Health centers in Niamey. Data were collected for socio demographic characteristics using a structured questionnaire. Samples were tested using ELISA and qRT-PCR for anti-HEV IgM and RNA detection respectively. HEV associated risk factors were determined using multivariate logistic regression models.

**Results:** A total of 491 pregnant women were enrolled with mean age of 26.5 years. The prevalence of anti-HEV IgM antibodies was 17.1%, while HEV RNA was detected in 3.46% of participants, indicating recent and active HEV infection. Anti-HEV IgM seropositivity was more frequent among women aged 26–30 years and those in the third trimester of pregnancy. Gravidity was significantly associated with HEV RNA positivity (p = 0.032). In multivariable analysis, the use of well water was independently associated with HEV infection (aOR = 75.70; 95% CI: 1.23–4643.20; p = 0.039).

**Conclusion:** In view of the high seroprevalence of HEV among pregnant women, preventive measures including hygiene sensitization should be prioritized. Further studies should be conducted to assess the real impact of HEV infection on maternal and fetal health.

## Introduction

Hepatitis E virus (HEV) infection is an emerging disease and a major global public health concern worldwide [1]. Globally, it is estimated that approximately 20 million HEV infections occur each year, resulting in about 44,000 deaths related to this infection [2]. In Africa, HEV prevalence was estimated at 29.13% [3], which is higher than the overall global prevalence of 16.5% [4]. In highly endemic regions, HEV is mainly transmitted via the fecal-oral route, through consumption of contaminated water, as well as by consumption of raw or undercooked meat from HEV-infected animals [5,6]. Phylogenetics characterization of the HEV identified eight distinct genotypes (HEV-1 to HEV-8) and 36 subtypes [7]. Human infection is mainly transmitted by HEV1 to 4, and a recently reported HEV-7 [8]. Usually, the infection is usually self-limited but may persist and cause chronic hepatitis in immunocompromised patients [9]. However, severe forms of the infection occur in different groups of vulnerable population including pregnant women [10]. Among this group, the case-fatality rates can be up to 40%, especially during the third quater of pregnancy [11]. In addition, serious maternal complications such as fulminant hepatic failure (FHF), eclampsia, preterm delivery, low birth weight, and stillbirth of the fœtus and postpartum hemorrhage can be observed [12,13]. In Niger Republic, a recent study reported a HEV prevalence of 32.12% among displaced population, with high death rate among pregnant women [8]. Therefore, it is essential to assess the prevalence of HEV among different populations, particularly asymptomatic pregnant women. We hereby aim to assess the prevalence of hepatitis E virus infection among pregnant women attending medical facilities in Niamey, the capital city of Niger.

## Materiel et methods

### 2.1 Ethical considerations

The proposal was approved by the Niger National Ethical Committee under number N°04/2024/CNERS as a less than minimal risk for research. Written consent was obtained from all pregnant women prior to sampling. All methods including the use of human samples were performed in accordance with the Declaration of Helsinki.

### 2.2 Study design and setting

The cross-sectional was conducted between February and May 2025. Pregnant women were enrolled at three maternity wards, namely Maternite Yantala, CSI Gamkalle and CSI Madina 2 located at different area in Niamey. These facilities provide prenatal care, delivery services, postnatal care, and various other health services. Pregnant women attend mainly these centers for antenatal consultations. A paper-based questionary was used to collect socio demographic and clinical data among consenting pregnant women.

### 2.3 Laboratory testing

#### 2.3.1. Sample collection and preparation

Blood sample was collected using 5 ml dry tube from each participant and transported to the Centre de Recherche Medicale et Sanitaire (CERMES) for laboratory analysis in accordance with triple-packaging transport procedures. After centrifugation at 3,000 rpm for 10 minutes, the serum was separated and stored in a −20 °C freezer prior to analysis.

#### 2.3.2. Anti-HEV IgM detection

The NovaLisa® Hepatitis E Virus (HEV) IgM (HEVM0780, Gold Standard Diagnostics, Germany) was used to detect anti-HEV IgM antibodies. According to the manufacturer, the test present high performance of 96.15% sensitivity and 98.59% specificity (14). Briefly, samples were added onto the pre-coated 96-well plates with specific antigens. After incubation and washing, Horseradish peroxidase (HRP) conjugated was added to the wells. Revelation of HEV IgM antibody was done by adding Tetramethylbenzidine (TMB) substrates and optical density was read using spectrophotometer at 450 nm with a selected reference wavelength within 620 nm. Positive and negative controls were included on each ELISA microplate. Samples with a value above the cutoff (> 11 NTU) were considered positive, indicating the detection of anti-HEV IgM antibodies.

#### 2.3.3. HEV RNA detection

All collected serum samples were analyzed by qRT-PCR for the detection of HEV RNA. Viral RNA was extracted from 200 μL of serum using the commercial Nucleic Acid Extraction Rapid Kit and the extraction was performed on the automated Bioperfectus GenMag system (Beijing, China) according to the manufacturer’s instructions [15]. Five microliters of the extracted RNA were used for HEV detection using the AgPath-ID™ One-Step RT-PCR Kit. Each 25 μL reaction mixture contained 12.5 μL of RT-PCR buffer, 5.5 μL of nuclease-free water, 0.5 μL of enzyme mix and primers and probe supplied [8]. Reverse transcription was performed at 45°C for 10 minutes, followed by an initial denaturation step at 95°C for 10 minutes. Amplification was then carried out over 45 cycles consisting of 95°C for 15 seconds and 55°C for 60 seconds. Real-time RT-PCR assays were performed using the QuantStudio™ FastStudio 5 Real-Time PCR System (Thermo Fisher Scientific, Waltham, MA, USA).

### 2.4. Data analysis

Data were analyzed using RStudio software (version 2025.05). Associations between hepatitis E positivity and explanatory variables were initially explored using bivariate analysis. A multivariable logistic regression model was then fitted to identify factors independently associated with hepatitis E seropositivity, with results reported as odds ratios (ORs) and their corresponding 95% confidence intervals (95% CIs). Statistical significance was assessed using a two-sided p-value < 0.05.

## Results

A total of 491 pregnant women included in the study were screened for the presence of anti-HEV IgM antibodies and HEV RNA. The study population was relatively young and their ages ranging from 13 to 45 years, with a mean age of 26.5 ± 6.3 years. The mean maternel age of the participants enrolled was 24.89± 8.43 weeks and 132 had experienced at least one episode of abortion, representing 26.9%.

Among the 491 pregnant women enrolled, 84 (17.1%) were positive for anti-HEV IgM antibodies, and 17 (3.46%) had detectable HEV RNA by qRT-PCR. Of the 17 HEV RNA-positive samples, 14 (82.35%) were also positive for anti-HEV IgM by ELISA, whereas three sample was positive for HEV RNA but negative for anti-HEV IgM antibodies (Figure 1).

**Figure 1:**
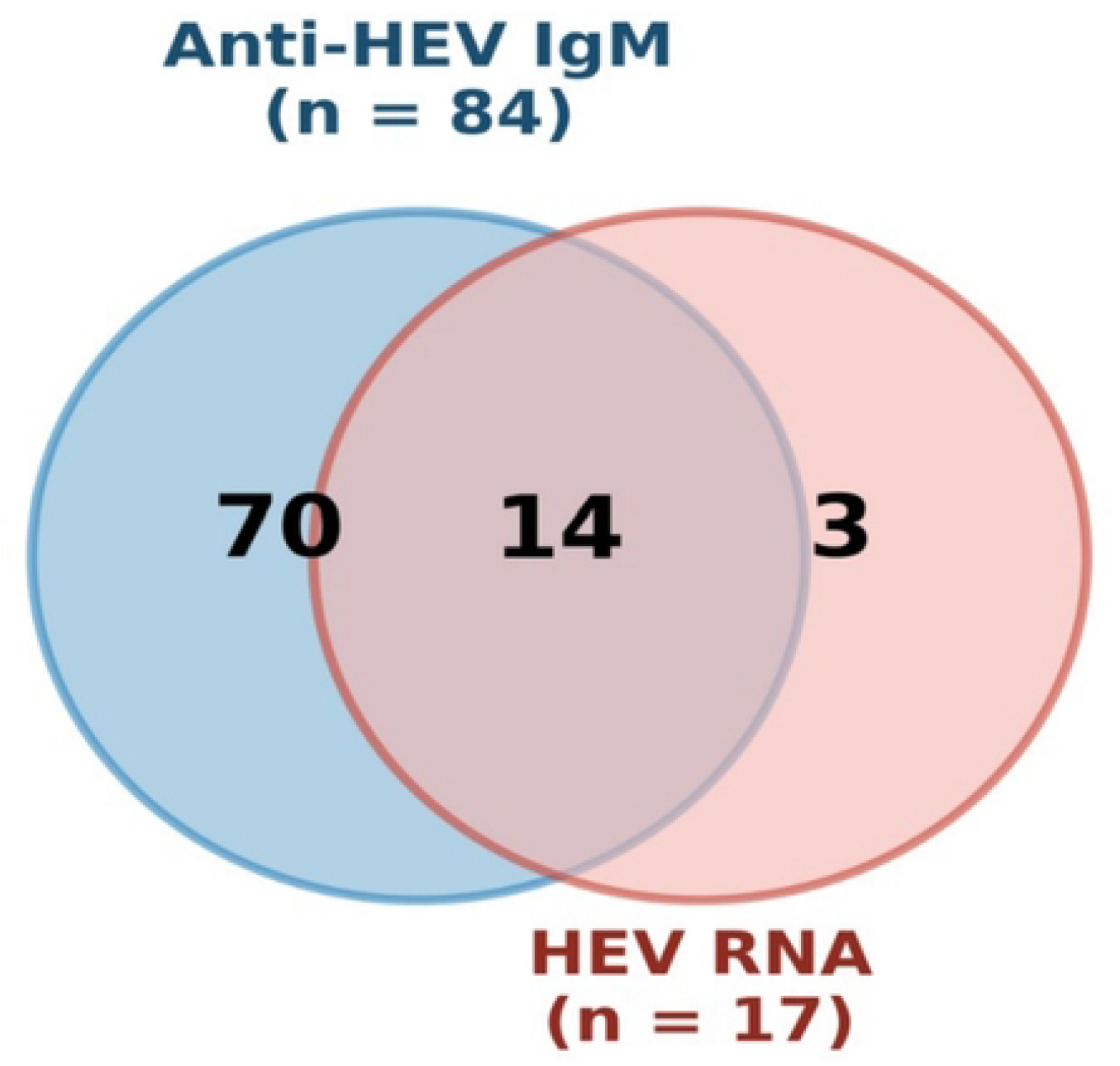
Concordance between anti-HEV IgM seropositivity and HEV RNA detection among pregnant women.

Women aged 26–30 years accounted for the highest proportion of positive cases (27.4%), followed by those aged 21–25 years (26.2%). Participants with no formal education represented 28.6% of the IgM-positive cases, whereas 44.0% had a secondary level of education. In addition, 72.6% of seropositive women lived in monogamous households, and 66.7% were housewives. Regarding obstetric characteristics, women in the second and third trimesters of pregnancy accounted for 45.2% and 46.4% of IgM-positive cases, respectively. Multiparous women represented 53.6% of seropositive participants, while nulliparous and primiparous women accounted for 29.8% and 16.7%, respectively. Grand multiparous women (22.6%) and multigravidae (22.6%) were the most represented categories among positive participants. None of the variables examined was significantly associated with anti-HEV IgM positivity *p* > 0.05. For the detection of viral RNA by qRT-PCR, a statistically significant association was observed only with gravidity (*p* = 0.032). Multigravidae accounted for 41.2% of qRT-PCR-positive cases, followed by primigravidae (29.4%) and grand multigravidae (23.5%). No significant association was found between qRT-PCR positivity and sociodemographics characteristics *p* > 0.05 (Table 1).

**Table 1:** Association between characteristics and anti-HEV IgM/hepatitis E virus RNA status.

| Characteristics | IgM HEV |  |  | p-value | qRT-PCR (RNA) |  | p-value |
| --- | --- | --- | --- | --- | --- | --- | --- |
|  | All (n=491) | Positive (n=84) | Negative (n=407) |  | Positive (n=17) | Negative (n=474) |  |
| **Age** |  |  |  | **0.596** |  |  | **0.290** |
| ≤20 (ref) | 101 (20.6) | 19 (22.6) | 82 (20.1) |  | 4 (23.5) | 97 (20.5) |  |
| 21-25 | 154 (31.4) | 22 (26.2) | 132 (32.4) |  | 2 (11.8) | 152 (32.1) |  |
| 26-30 | 109 (22.2) | 23 (27.4) | 86 (21.1) |  | 4 (23.5) | 105 (22.2) |  |
| 31-35 | 56 (11.4) | 10 (11.9) | 46 (11.3) |  | 2 (11.8) | 54 (11.4) |  |
| ≥26 | 71 (14.5) | 10 (11.9) | 61 (15.0) |  | 5 (29.4) | 66 (13.9) |  |
| **Formal education** |  |  |  | **0.802** |  |  | **0.209** |
| No formal education | **107 (21.8)** | **24 (28.6)** | **83 (20.4)** |  | **7 (41.2)** | **100 (21.1)** |  |
| Primary | 90 (18.3) | 13 (15.5) | 77 (18.9) |  | 4 (23.5) | 86 (18.1) |  |
| Secondary | 222 (45.2) | 37 (44.0) | 185 (45.5) |  | 6 (35.5) | 216 (45.6) |  |
| Tertiary | 72 (14.7) | 10 (11.9) | 62 (15.2) |  | 0 (0.0) | 72 (15.2) |  |
| No formal education | 107 (21.8) | 24 (28.6) | 83 (20.4) |  | 7 (41.2) | 100 (21.1) |  |
| **Type of family** |  |  |  | **0.588** |  |  | **0.151** |
| Monogamous | 368 (74.9) | 61 (72.6) | 307 (75.4) |  | 10 (58.8) | 358 (75.5) |  |
| Polygamous | 123 (25.1) | 23 (27.4) | 100 (24.6) |  | 7 (41.2) | 116 (24.5) |  |
| **Occupation** |  |  |  | **0.582** |  |  | **0.297** |
| Government (ref) | 30 (6.1) | 3 (3.6) | 27 (6.6) |  | 0 (0.0) | 30 (6.3) |  |
| Housewife | 332 (67.6) | 56 (66.7) | 276 (67.8) |  | 14 (82.4) | 318 (67.1) |  |
| Self employed | 74 (15.1) | 13 (15.5) | 61 (15.0) |  | 3 (17.6) | 71 (15.0) |  |
| Student | 55 (11.2) | 12 (14.3) | 43 (10.6) |  | 0 (0.0) | 55 (11.6) |  |
| **Number of pregnancy trimester** |  |  |  | **0.670** |  |  | **0.649** |
| First trimester (ref) | 50 (10.2) | 7 (8.3) | 43 (10.6) |  | 1 (5.9) | 49 (10.3) |  |
| Second trimester | 203 (41.3) | 38 (45.2) | 165 (40.5) |  | 6 (35.3) | 197 (41.6) |  |
| Third trimester | 238 (48.5) | 39 (46.4) | 199 (48.9) |  | 10 (58.8) | 228 (48.1) |  |
| **Parity** |  |  |  | **0.236** |  |  | **0.218** |
| Multiparous (ref) | 252 (51.3) | 45 (53.6) | 207 (50.9) |  | 11 (64.7) | 241 (50.8) |  |
| Primaparous | 115 (23.4) | 14 (16.7) | 101 (24.3) |  | 1 (5.9) | 119 (25.1) |  |
| Nulliparous | 124 (25.4) | 25 (29.8) | 99 (24.8) |  | 5 (29.4) | 114 (24.1) |  |
| **Gravidity** |  |  |  | **0.696** |  |  | **0.032\*** |
| Grand multipara | 95 (19.3) | 19 (22.6) | 76 (18.7) |  | 4 (23.5) | 91 (19.2) |  |
| Multigravida | 108 (22.0) | 19 (22.6) | 89 (21.9) |  | 7 (41.2) | 101 (21.3) |  |
| Paucigravida | 190 (38.7) | 28 (33.3) | 162 (39.8) |  | 1 (5.9) | 189 (39.9) |  |
| Primigravida | 98 (20.0) | 18 (21.4) | 80 (19.7) |  | 5 (29.4) | 93 (19.6) |  |
| **Habitation** |  |  |  | **0.655** |  |  | **0.274** |
| Rural | 9 (1.8) | 2 (2.4) | 7 (1.7) |  | 1 (5.9) | 8 (1.7) |  |
| Urban | 482 (98.2) | 82 (97.6) | 400 (98.3) |  | 16 (94.1) | 466 (98.3) |  |

After adjustment for the variables included in the model, only the source of drinking water remained significantly associated with positivity. Patients using well water had a significantly higher probability of positivity (adjusted OR [aOR] = 75.70; 95% CI: 1.23–4643.20; p = 0.039) compared to those using tap and water tower. No statistically significant association was observed between positivity and handwashing practices, pet ownership of domestic animals and educational level (all *p* > 0.05). (Table 2)

**Table 2:** Multivariate analysis between infection of HEV and risk factors.

| Variable /<br>modalité | Mesure ajustée | IC95% | p |
| --- | --- | --- | --- |
| **Handwashing** |  |  |  |
| *No (référence)* | *1,00 (réf.)* | — | — |
| Yes | ORa = 0,29 | [0,05–1,82] | 0,189 |
| **Pet ownership** |  |  |  |
| *No (référence)* | *1,00 (réf.)* | — | — |
| Yes | ORa = 0,00 | [0–] | 1,000 |
| **Water source used** |  |  |  |
| *Tap water*<br>*(référence)* | *1,00 (réf.)* | — | — |
| Water tower | ORa = 0,58 | [0,11–2,94] | 0,508 |
| Water well | ORa = 50,92 | [1,40–1847,02] | 0,032 |
| **Formal education** |  |  |  |
| *No formal*<br>*education*<br>*(référence)* | *1,00 (réf.)* | — | — |
| Primary | ORa = 0,87 | [0,23–3,28] | 0,833 |
| Secondary | ORa = 0,43 | [0,11–1,67] | 0,224 |
| Tertiary | ORa = 0,00 | [0–] | 1,000 |

## Discussion

Hepatitis E virus is a major cause of liver disease worldwide and, although long-term sequelae are rare, the infection is associated with substantial mortality among pregnant women [15]. In our study, the positivity rate of HEV infection was 17.1% for anti-HEV IgM and 3.46% for HEV RNA, suggesting active circulation of the virus and the presence of recent infections within the study population. This seroprevalence is comparable to those reported in Gabon (14.1%), Nigeria (13%), and Cameroon (13%) [12,16,17], suggesting that HEV remains an important public health concern across several sub-Saharan African countries. In contrast, it is substantially higher than the estimates reported in Benin, Ethiopia, and Senegal (0.5–1.44%), but considerably lower than that observed in Bangladesh (55.3%) [18–21]. These differences probably reflect variations in epidemiological settings, including access to safe drinking water, sanitation conditions, the endemicity of HEV, as well as differences in study populations and diagnostic approaches.

Beyond the overall prevalence, the distribution of recent infections according to participants’ characteristics provides further insight into the epidemiology of HEV infection. Most recent infections occurred among women aged 21–30 years, consistent with the findings reported by Diaoura et al. in Senegal [20]. However, no significant association was observed between age and anti-HEV IgM positivity. This distribution is therefore likely to reflect the demographic structure of the study population, in which women within this age group represented the majority of participants, rather than an age-related increase in susceptibility.

Similarly, the distribution of HEV infection according to obstetric characteristics revealed several noteworthy patterns. The highest prevalence of anti-HEV IgM antibodies was observed among women in the third trimester of pregnancy. This finding is consistent with that reported by Kumar [22], who showed that most HEV-infected pregnant women were in the third trimester of pregnancy. Previous studies have demonstrated that HEV infection during pregnancy, particularly during the second and third trimesters, may lead to acute liver failure and substantially increase the risk of maternal mortality [12,23,24]. Nevertheless, no statistically significant association was found between gestational trimester and anti-HEV IgM positivity in our study, suggesting that the risk of recent HEV infection was not influenced by gestational age in this population.

In contrast to gestational age, a statistically significant association was observed between HEV infection and gravidity. This association may be explained by the fact that multigravid women, owing to a greater number of pregnancies, may have experienced more frequent exposure to HEV throughout their reproductive years, thereby increasing their likelihood of recent infection. In addition, repeated pregnancies are often associated with lower socioeconomic conditions, larger household sizes, and more frequent exposure to environments with inadequate hygiene and sanitation, all of which may facilitate the fecal–oral transmission of HEV [25]. These findings underscore the need for strengthened surveillance of pregnant women, particularly considering that during the 2017 HEV outbreak in Niger, 19 of the 42 reported deaths (45.23%) occurred among pregnant women [10].

Socioeconomic factors also appeared to play an important role in HEV infection in our study population. Previous studies have consistently demonstrated the protective effect of education, with significantly lower HEV seroprevalence among educated women [26]. These observations are consistent with our findings. Women with a university education or higher had the lowest HEV seropositivity among all participants. Moreover, pregnant women with no formal education had approximately twice the risk of HEV infection compared with those with a university education or higher. This difference may be explained by lower health literacy, which influences knowledge and the adoption of appropriate personal, food, and environmental hygiene practices [27]. Consistent with this interpretation, multivariable analysis also demonstrated a significant association between HEV infection and the source of drinking water, further highlighting the central role of water quality in the fecal–oral transmission of HEV. These socioeconomic determinants may also partly explain the relatively high prevalence observed in our study. Samples were collected exclusively from public healthcare facilities, where services are provided free of charge or at very low cost. These facilities are more frequently attended by women from socioeconomically disadvantaged backgrounds. Consequently, our study population may not be representative of pregnant women with higher socioeconomic and educational status.

Our study has some limitations. All study sites were located in urban hospital settings, which may limit the generalizability of our findings to pregnant women living in rural areas or those who do not attend antenatal care. Anti-HEV IgG antibodies were not assessed, preventing the estimation of the proportion of pregnant women who had been previously exposed to the hepatitis E virus. In addition, pregnant women who tested positive for anti-HEV IgM antibodies were not followed prospectively, precluding the assessment of the maternal and neonatal outcomes of HEV infection, particularly its impact on newborns.

## Conclusion

This study demonstrates active circulation of hepatitis E virus among pregnant women in Niamey, with a high prevalence of recent HEV infection. The association between HEV infection and the use of well water highlights the importance of improving access to safe drinking water and sanitation to reduce fecal–oral transmission. Given the increased risk of severe maternal outcomes associated with HEV infection during pregnancy, strengthening surveillance, integrating HEV screening into antenatal care in high-risk settings, and implementing preventive public health measures should be considered. Further prospective studies are needed to assess maternal and neonatal outcomes and to better characterize the molecular epidemiology of HEV in Niger.

## Data Availability

The minimal data is available at Fatimata Hassane and Adamou Lagare, the corresponding authors.

## Acknowledgments

We acknowledge the efforts of all cited authors whose works contributed to the development of this paper.

## Supporting information

S1 Figure 1: Concordance between anti-HEV IgM seropositivity and HEV RNA detection among pregnant women

## Références bibliographiques

1. Wu C, Wu X, Xia J. Hepatitis E virus infection during pregnancy. Virol J. 10 juin 2020;17:73. doi:10.1186/s12985-020-01343-9 PubMed PMID: 32522266; PubMed Central PMCID: PMC7286216.

2. Ismail MB, Khodor S, Osman M, Mallat H, Dabboussi F, Hamze M. Seroprevalence of hepatitis E virus in pregnant women in northern Lebanon. East Mediterr Health J. 1 mai 2020;26(5):580–5. doi:10.26719/emhj.19.103

3. Dagnew M, Belachew A, Tiruneh M, Moges F. Hepatitis E virus infection among pregnant women in Africa: systematic review and meta-analysis. BMC Infect Dis. 13 juin 2019;19:519. doi:10.1186/s12879-019-4125-x PubMed PMID: 31195988; PubMed Central PMCID: PMC6567642.

4. Ahmad T, Hui J, Musa TH, Behzadifar M, Baig M. Seroprevalence of hepatitis E virus infection in pregnant women: a systematic review and meta-analysis. Ann Saudi Med. mars 2020;40(2):136–46. doi:10.5144/0256-4947.2020.136 PubMed PMID: 32241162; PubMed Central PMCID: PMC7118233.

5. Sclair SN, Schiff ER. An Update on the Hepatitis E Virus. Curr Gastroenterol Rep. 12 janv 2013;15(2):304. doi:10.1007/s11894-012-0304-2

6. Wakabayashi Y, Kitaura S, Okamoto K, Ikeda M, Yanagimoto S, Okugawa S, et al. Hepatitis E virus seroprevalence in patients with human immunodeficiency virus: A single-center study in Japan. J Clin Virol Plus. août 2024;4(3):100185. doi:10.1016/j.jcvp.2024.100185

7. Sooryanarain H, Meng XJ. Hepatitis E virus: Reasons for emergence in humans. Curr Opin Virol. févr 2019;34:10–7. doi:10.1016/j.coviro.2018.11.006 PubMed PMID: 30497051; PubMed Central PMCID: PMC6476702.

8. Lagare A, Abdoulkader IK, Fall G, Ousmane H, Hounkanrin W, Aoula B, et al. Molecular epidemiology of Hepatitis E virus among humans in the Niger Republic, 2017–2023. J Clin Virol. févr 2025;176:105761. doi:10.1016/j.jcv.2025.105761

9. Luo Q, Chen J, Zhang Y, Xu W, Liu Y, Xie C, et al. Viral hepatitis E: Clinical manifestations, treatment, and prevention. Liver Res. mars 2024;8(1):11–21. doi:10.1016/j.livres.2024.01.001

10. Lagare A, Ibrahim A, Ousmane S, Issaka B, Zaneidou M, Kadadé G, et al. Outbreak of Hepatitis E Virus Infection in Displaced Persons Camps in Diffa Region, Niger, 2017. Am J Trop Med Hyg. oct 2018;99(4):1055–7. doi:10.4269/ajtmh.17-0950 PubMed PMID: 30062983; PubMed Central PMCID: PMC6159588.

11. N K, J I, N P, R A, A L, H W, et al. Hepatitis E virus infection. Nat Rev Dis Primer. 16 nov 2017;3. doi:10.1038/nrdp.2017.86 PubMed PMID: 29154369.

12. Ajayi B, Igwegbe I, Latbone S, Oderinde B, Kida I, Oyinloye S, et al. Seroprevalence and associated risk factors of hepatitis E virus infection among pregnant women attending Maiduguri teaching hospital, Nigeria. Microbes Infect Dis. 30 août 2020;0(0):0–0. doi:10.21608/mid.2020.39788.1053

13. Ms K, s K, Ms K. Clinical course and duration of viremia in vertically transmitted hepatitis E virus (HEV) infection in babies born to HEV-infected mothers. J Viral Hepat. juill 2009;16(7). doi:10.1111/j.1365-2893.2009.01101.x PubMed PMID: 19228284.

14. brochure_gsd-novalisageneral_0522_en.pdf.

15. Nucleic Acid Extraction Rapid Kit (Magnetic Bead Method) | Bioperfectus [Internet]. [cité 4 août 2026]. Disponible sur: https://www.bioperfectus.com/productdetail/nucleicacidextractionrapidkitmagneticbeadmethod

16. Teo CG. Fatal outbreaks of jaundice in pregnancy and the epidemic history of hepatitis E. Epidemiol Infect. mai 2012;140(5):767–87. doi:10.1017/S0950268811002925

17. Caron M, Kazanji M. Hepatitis E virus is highly prevalent among pregnant women in Gabon, central Africa, with different patterns between rural and urban areas. Virol J. 22 éc 2008;5:158. doi:10.1186/1743-422X-5-158 PubMed PMID: 19102767; PubMed Central PMCID: PMC2628354.

18. Af M, M AA, Cg M, M N, R N. Seroprevalence of hepatitis E virus antibodies in different human populations of Cameroon. J Med Virol. nov 2019;91(11). doi:10.1002/jmv.25545 PubMed PMID: 31297845.

19. De Paschale M, Ceriani C, Romanò L, Cerulli T, Cagnin D, Cavallari S, et al. Epidemiology of hepatitis E virus infection during pregnancy in Benin. Trop Med Int Health. 2016;21(1):108–13. doi:10.1111/tmi.12632

20. Niguse S, Hailekiros H, Buruh G, Dejene T, Berhe N, Asmelash T. Seroprevalence and risk factors of Hepatitis E virus infection among pregnant women attending antenatal care in health facilities of Tigray, Northern Ethiopia. J Med Virol. août 2018;90(8):1364–9. doi:10.1002/jmv.25190

21. Diouara AAM, Lo S, Nguer CM, Senghor A, Diop Ndiaye H, Manga NM, et al. Hepatitis E Virus Seroprevalence and Associated Risk Factors in Pregnant Women Attending Antenatal Consultations in Senegal. Viruses. 9 août 2022;14(8):1742. doi:10.3390/v14081742 PubMed PMID: 36016364; PubMed Central PMCID: PMC9416362.

22. Sultana R, Rozhana S, Tabassum S, Munshi SU. Feto-Maternal Outcomes of Hepatitis E Virus Infection in Pregnancy of Bangladeshi women. Bangladesh J Med Microbiol. 13 févr 2017;9(2):3–6. doi:10.3329/bjmm.v9i2.31413

23. Kumar S, Subhadra S, Singh B, Panda BK. Hepatitis E virus: the current scenario. Int J Infect Dis. 1 avr 2013;17(4):e228–33. doi:10.1016/j.ijid.2012.11.026 PubMed PMID: 23313154.

24. Chaudhry SA, Verma N, Koren G. Hepatitis E infection during pregnancy. Can Fam Physician. juill 2015;61(7):607-8. PubMed PMID: 26175368; PubMed Central PMCID: PMC4501603.

25. Ehi Airiohuodion P, Wartel A, Yako AB, Mac PA. Seroprevalence and burden of hepatitis E viral infection among pregnant women in central Nigeria attending antenatal clinic at a Federal Medical Centre in Central Nigeria. Front Med. 2 sept 2022;9:888218. doi:10.3389/fmed.2022.888218 PubMed PMID: 36117965; PubMed Central PMCID: PMC9478474.

26. Koyuncu A, Mapemba D, Ciglenecki I, Gurley ES, Azman AS. Setting a Course for Preventing Hepatitis E in Low and Lower-Middle-Income Countries: A Systematic Review of Burden and Risk Factors. Open Forum Infect Dis. 13 avr 2021;8(6):ofab178. doi:10.1093/ofid/ofab178 PubMed PMID: 34113684; PubMed Central PMCID: PMC8186248.

27. Begum N, Devi SG, Husain SA, Ashok Kumar, Kar P. Seroprevalence of subclinical HEV infection in pregnant women from north India: a hospital based study. Indian J Med Res. éc 2009;130(6):709-13. PubMed PMID: 20090131.

28. Ike W. Seroprevalence of hepatitis-E virus infection among pregnant women at a secondary healthcare facility in Abeokuta, Southwest, Nigeria. Niger J Pharm. 30 mars 2024;58(1). doi:10.51412/psnnjp.2024.09

